# Facilitators and barriers to community pharmacy PrEP delivery: A scoping review

**DOI:** 10.1101/2023.10.06.23296672

**Authors:** China Harrison, Hannah Family, Joanna Kesten, Sarah Denford, Anne Scott, Sarah Dawson, Jenny Scott, Caroline Sabin, Joanna Copping, Lindsey Harryman, Sarah Cochrane, Jeremy Horwood

**Author notes:** Corresponding author: China Harrison https://orcid.org/0000-0003-3671-7866.

## Abstract

**Introduction:** Pre-exposure prophylaxis (PrEP) is an effective medication to reduce the risk of acquiring HIV. PrEP is available free of charge in the United Kingdom from sexual health clinics. Expanding PrEP delivery to community pharmacies holds promise and aligns with UK government goals to eliminate new cases of HIV by 2030. The aim of this scoping review was to describe the existing evidence about the barriers to and facilitators of community pharmacy PrEP delivery, for pharmacists and pharmacy clients, as aligned with the Capacity Opportunity, Motivation Behaviour (COM-B) Model.

**Methods:** Five bibliographic and five review databases were searched from inception to August 2023. Literature of any study design was included if it discussed barriers and facilitators of community pharmacy PrEP delivery. Trial registrations, protocols and news articles were excluded.

**Results:** A total of 649 records were identified, 73 full texts were reviewed, and 56 met the inclusion criteria. Most of the included literature was original research (55%), from the United States of America (77%) conducted during or after the year 2020 (63%). Barriers to PrEP delivery for pharmacists included lack of knowledge, training and skills (capability), not having the necessary facilities (opportunity), concern about the costs of PrEP and believing that PrEP use could lead to risk behaviour’s and STIs (motivation). Facilitators included staff training (capability), time, the right facilities (opportunity), believing PrEP could be a source of profit and could reduce new HIV infections (motivation). For clients, barriers included lack of PrEP awareness (capability), pharmacy facilities (opportunity) and not considering pharmacists as healthcare providers (motivation). Facilitators included awareness of PrEP and pharmacist’s training to deliver it (capability), the accessibility of pharmacies (opportunity) and having an interest in PrEP (motivation).

**Conclusion:** To effectively enhance PrEP delivery in UK community pharmacies, the identified barriers and facilitators should be explored for relevance in the UK and subsequently addressed and leveraged at the pharmacy team, client and care pathway level. By comprehensively considering all aspects of the COM-B framework, community pharmacies could become crucial providers in expanding PrEP accessibility, contributing significantly to HIV prevention efforts.

## Introduction

The Human immunodeficiency Virus (HIV) epidemic remains a significant public health concern worldwide and the United Kingdom (UK) is no exception. In 2021, it was estimated that there were 105,200 people living with HIV in the UK, approximately 2,955 of whom had been newly diagnosed in that year (1, 2).

In recent years, Pre-Exposure Prophylaxis (PrEP) has emerged as an effective preventative biomedical intervention which reduces the risk of acquiring HIV by 92-99% (3–7). PrEP can be taken orally daily or ‘on demand’ before and after sex. To initiate PrEP, eligible clients are required to have baseline and regular health checks to ensure client safety. These include kidney function tests and sexually transmitted infections (STI) screening (8). Whilst internationally PrEP is available via pharmacies in some countries, in the UK it is only available free of charge via National Health Service (NHS) sexual health clinics.

Uptake of PrEP has been slow in some regions and among some demographic groups. Disproportionately low uptake has been observed among some populations at elevated risk of acquiring HIV, such as trans individuals, cisgender women, young people (9) and people of Black African or Caribbean origin (10, 11). Low uptake has been attributed to several barriers in accessing PrEP, including the geographical proximity of PrEP providers, stigma, and lack of awareness of PrEP (12) with 79% of people in need of PrEP, having had their need identified during a clinical consultation. This highlights the need to raise awareness of PrEP to support individuals’ treatment seeking autonomy (1, 12). Expanding PrEP delivery to community pharmacies could be an effective way to reduce barriers to access, enhance autonomy and increase the utilisation and coverage of this effective HIV prevention (2, 7).

Community pharmacies play a crucial role in delivering public health services and have expanding roles in health promotion and prescribing (13, 14). Predominantly they are private businesses where pharmacists and their teams use their expert knowledge to clinically screen and dispense prescriptions, sell or supply over the counter medicines, give advice and deliver locally commissioned healthcare services. Their accessibility, convenience, and customer rapport make them well-suited to address urgent and preventative care needs (15, 16). PrEP delivery could align well with existing service provision within pharmacies with pharmacy teams routinely dispensing prescriptions for sexual and reproductive health medicines (e.g. emergency hormonal contraception). Further, pharmacy-based interventions have demonstrated success in improving medication adherence for various medications including PrEP (17). For example, in the United States of America (USA), a pharmacy in Seattle implemented the One Step PrEP program, enabling pharmacists to prescribe and manage PrEP which resulted in a 90% PrEP adherence rate among attendees who had two or more visits (18). While implementing community pharmacy PrEP delivery in the UK could greatly benefit individuals at risk of HIV, community pharmacy PrEP delivery would require training, support and a behavioural change by pharmacists and people (hereafter, clients) attending their services.

The Capability Opportunity Motivation Behaviour (COM-B) model offers a valuable framework to understand the barriers and facilitators associated with community pharmacy PrEP delivery (19). According to the COM-B model, behaviour is a result of an interaction between capability, opportunity and motivation. Capability refers to an individual’s psychological (knowledge) and physical (skills) ability to participate in an activity. Opportunity refers to external factors that can be social (societal influences) or environmental (physical) that make a behaviour possible, and motivation refers to the reflexive (beliefs, intentions) or automatic (emotion) cognitive processes that direct and inspire behaviour (19). By examining the interplay of capability, opportunity and motivation relating to PrEP delivery, we can gain insights into the acceptability and feasibility of implementing community pharmacy PrEP delivery in the UK. This could help inform the development of targeted interventions and strategies to optimise PrEP delivery, thus advancing HIV prevention efforts and improving health outcomes in the UK.

The objective of this scoping review was to map and describe the existing evidence about the barriers to and facilitators of community pharmacy PrEP delivery for pharmacists and clients, according to the COM-B Model.

## Method

A scoping review was conducted following the methodological framework of scoping reviews (21), in accordance with the PRISMA (Preferred Reporting Items for Systematic Reviews and Meta Analyses) extension for scoping reviews (22). We used the five stage scoping framework designed by Arksey and O’Malley (23) that involved 1) identifying the research question; 2) identifying relevant studies; 3) selecting studies; 4) charting the data; and 5) collating, summarising and reporting the results.

*Stage 1:* The research question, “What is known from the previous literature about the barriers to and facilitators of community pharmacy PrEP delivery?” guided this review.

*Stage 2:* We used search terms related to HIV, PrEP and community pharmacies (see Supplementary Table 1) to search five main bibliographic databases (MEDLINE (Ovid); Embase (Ovid); PsycINFO (Ovid); CINAHL (EBSCOhost); and the Cochrane Central Register of Controlled Trials (CENTRAL)) and five review databases (Cochrane Database of Systematic Reviews (CDSR); Database of Promoting Health Effectiveness Reviews (DoPHER); Epistemonikos; Health Evidence; and NIHR Health Technology Assessments) from inception to August 2023. A manual search of the reference list of included literature and reviews related to the topic was also conducted to identify additional eligible research.

Search results were imported into Endnote (24) and duplicates were removed. The final entries were then imported to Rayyan (25) and two authors (JH & AS or CH) independently screened each title, abstract and full text according to the predefined inclusion/exclusion criteria (see below). Throughout the screening process, differences in opinions were resolved through discussion.

*Stage 3:* Literature was considered if it: 1) explored community pharmacy PrEP delivery; 2) included community pharmacists, PrEP clients, stakeholders, analysed data on PrEP initiation, continuation and adherence or explored community pharmacy PrEP interventions; 3) included studies about encouragement of PrEP use (e.g., willingness, attitudes intentions); and 4) included information on the barriers to and facilitators of community pharmacy PrEP delivery. Trial registrations, protocols and news articles were excluded.

*Stage 4:* Data extraction was performed by CH, JH, JK, HF and SD and included author(s), year of publication, country, sample, study objective, methodology, and barriers and facilitators. Methodological quality assessment was not performed (26).

*Stage 5:* An overview of the volume and nature of the available evidence is represented graphically. The barriers to and facilitators of community pharmacy PrEP delivery are tabulated and synthesised according to the COM-B model for pharmacists and clients.

## Results

A total of 649 records were identified. After duplicate removal, the remaining 467 titles and abstract were screened, resulting in exclusion of 394 articles. The remaining 73 records underwent full text review, and 19 were excluded. From searching the references of the included studies, a further two studies were identified. A total of 56 records were included in the scoping review (see Figure 1).

**Figure 1.**
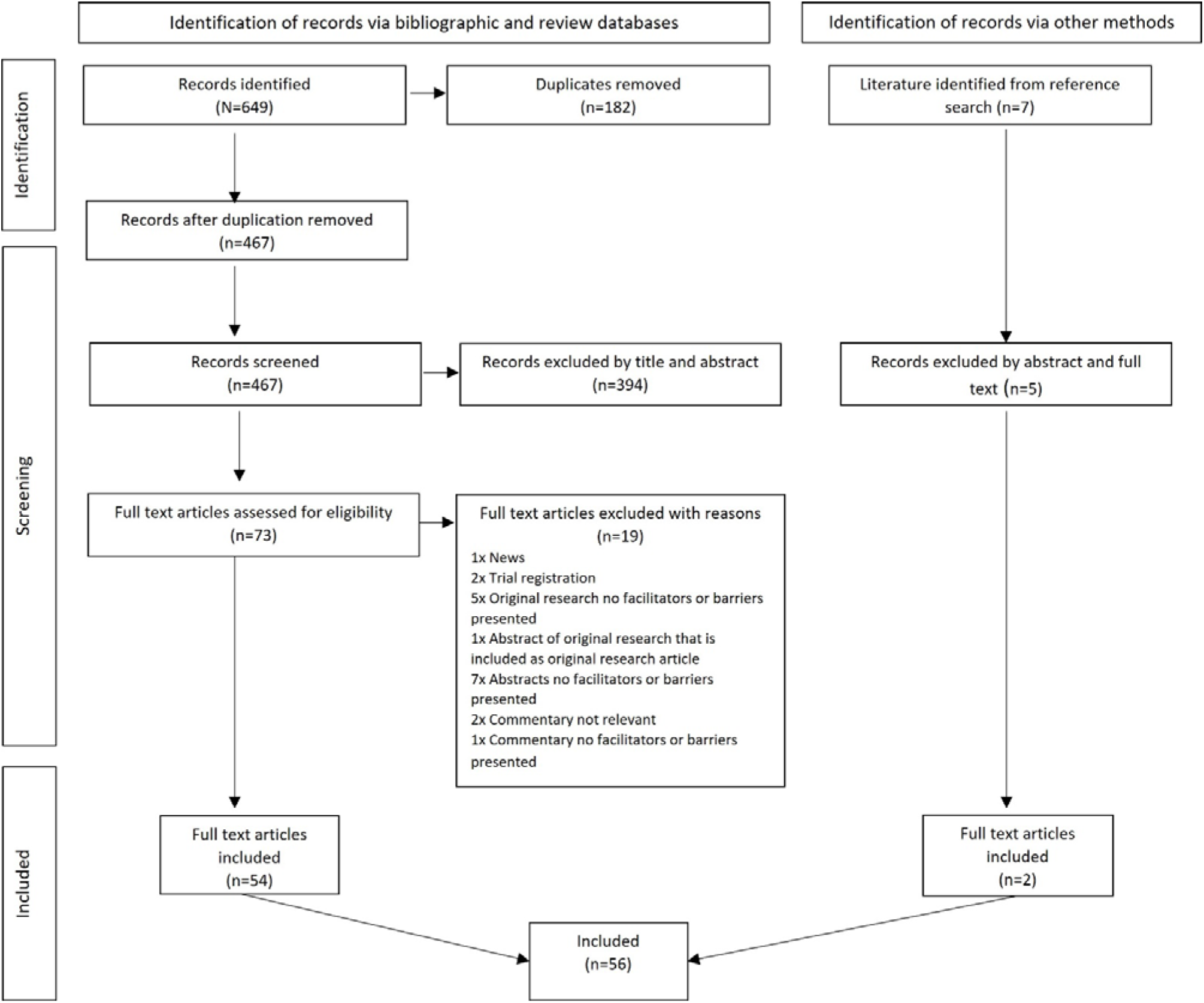
PRISMA flow diagram

As shown in figure 2, most of the included literature was original research (55%), commentaries (16%) or reviews (16%), from the USA (77%), and was conducted during or after 2020 (63%). The methodological characteristics, study objectives and population, of the 56 included records are summarised in Supplementary Table 2.

**Figure 2.**
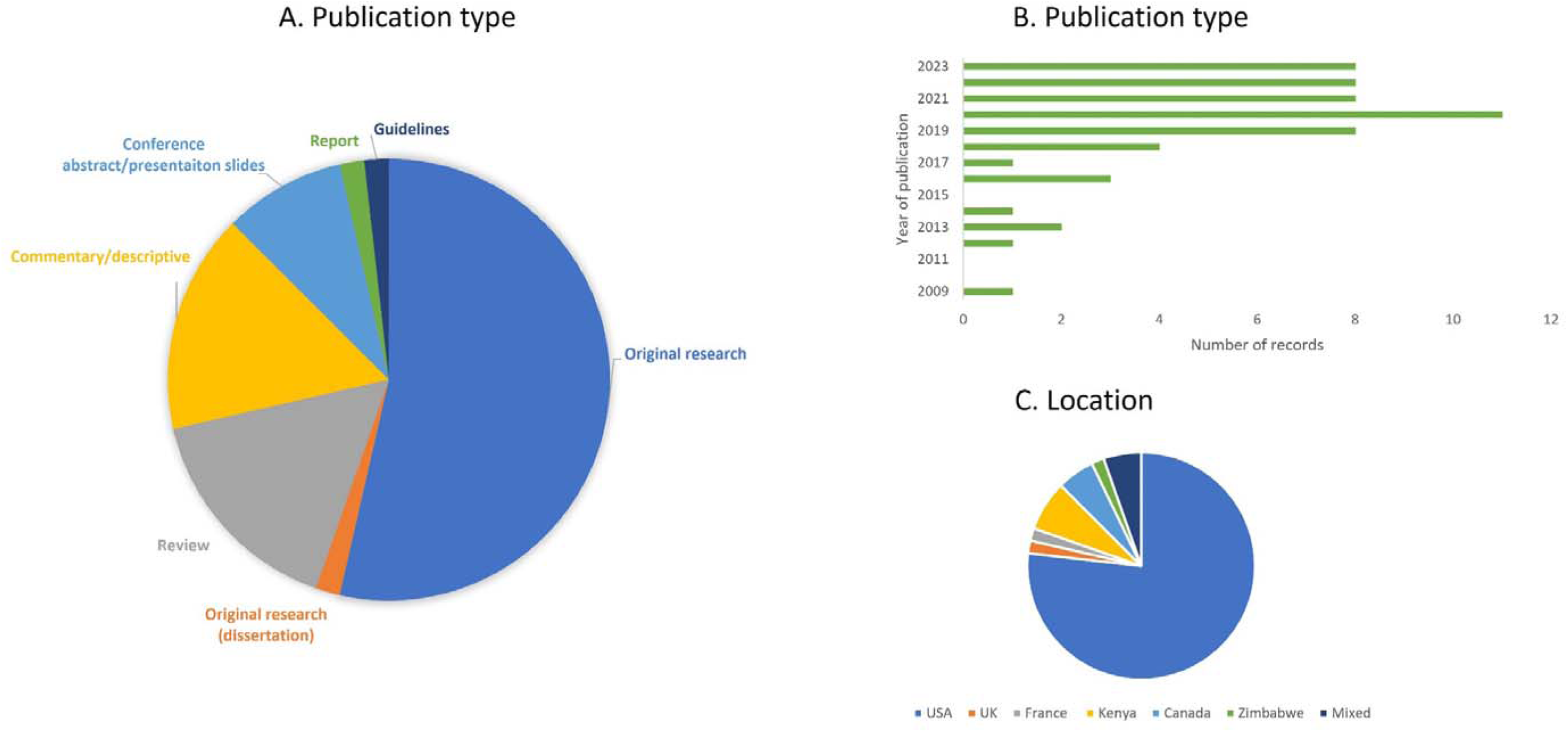
Characteristics of all the included records: Publication type (A), year of publication (B) and location (i.e., origin of study population or research) (C). Mixed location represents systematic reviews covering literature from various countries.

A summary of the barriers and facilitators, according to the COM-B model, are presented in Table 1 and synthesised descriptively below.

**Table 1.**
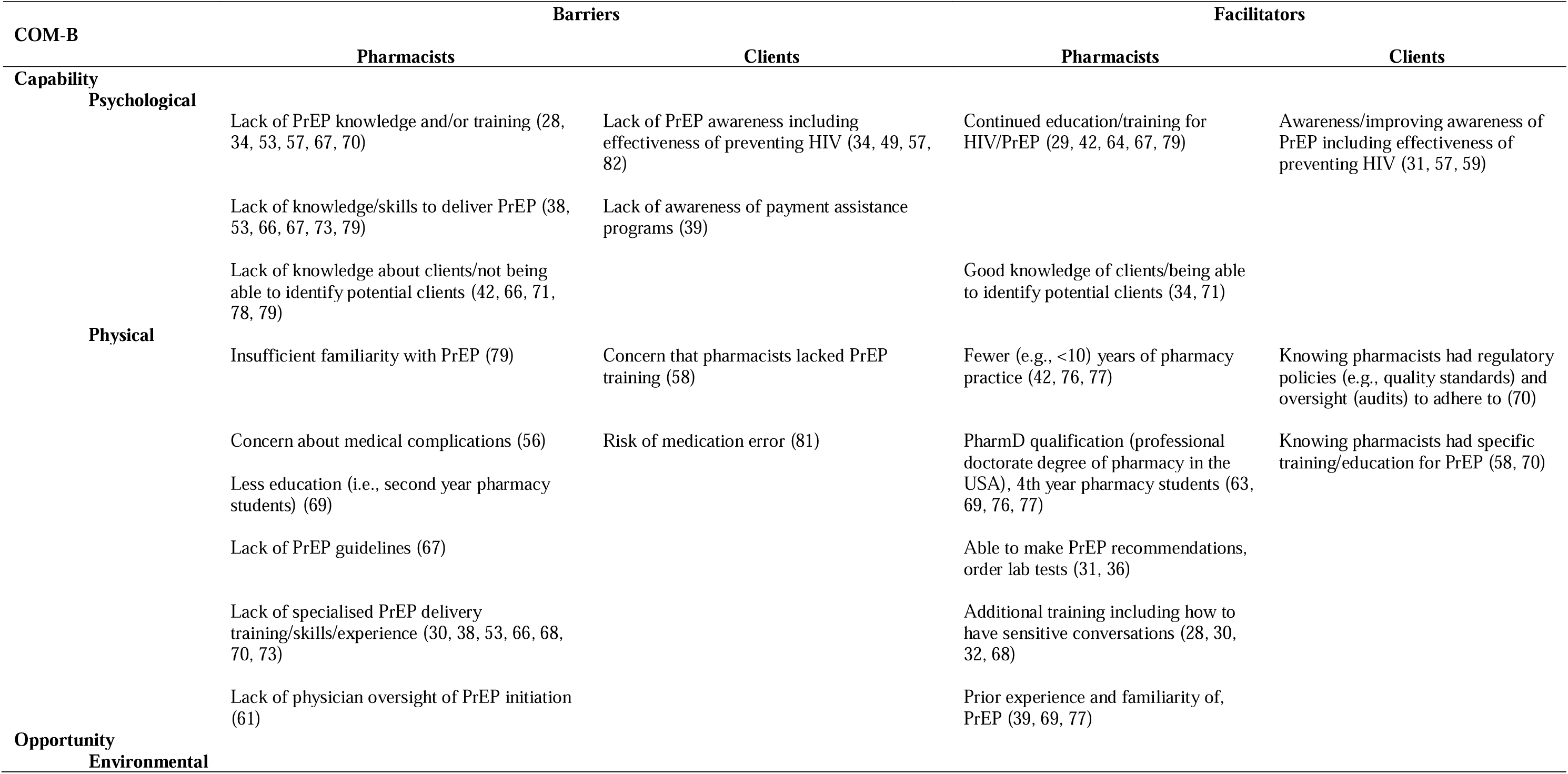
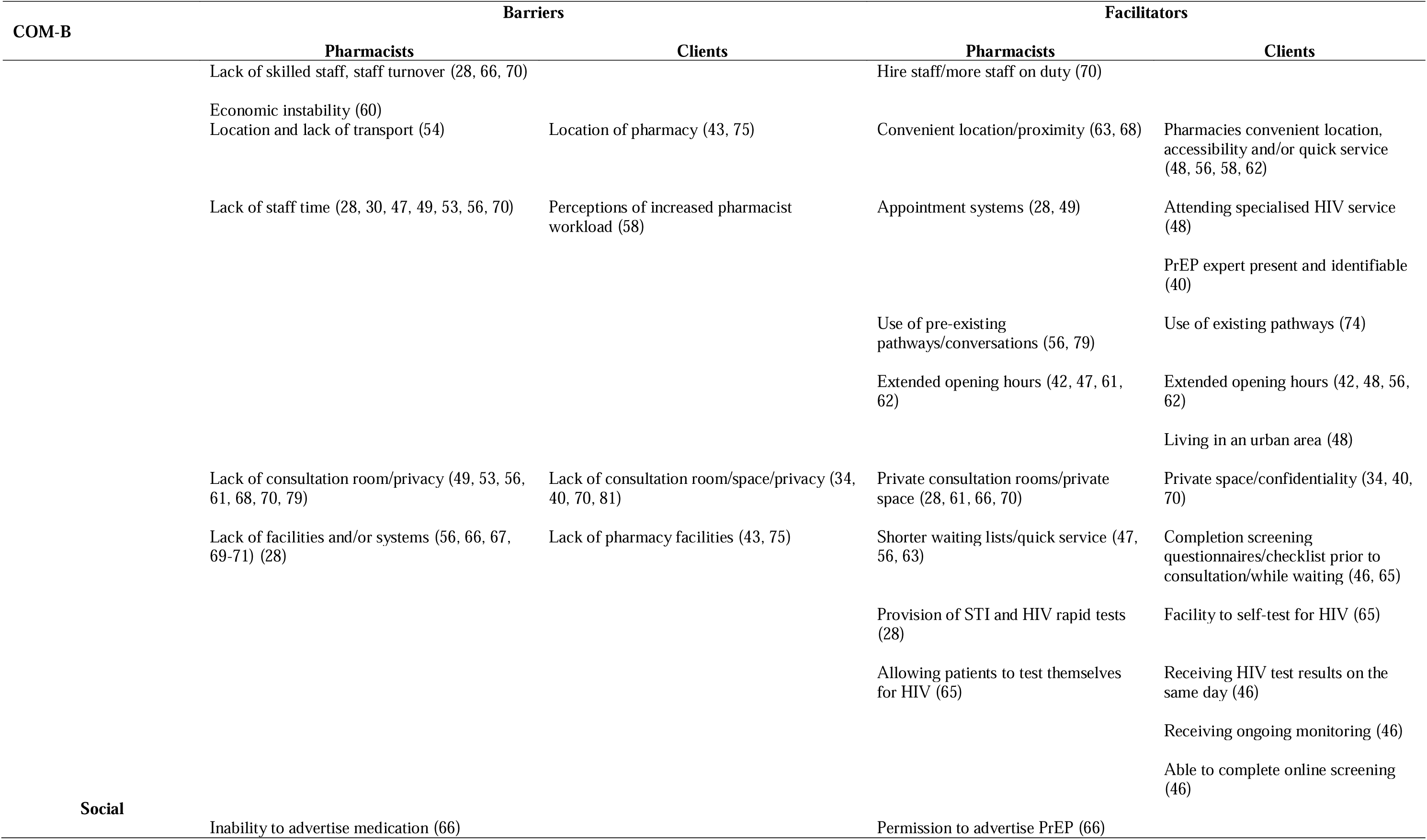
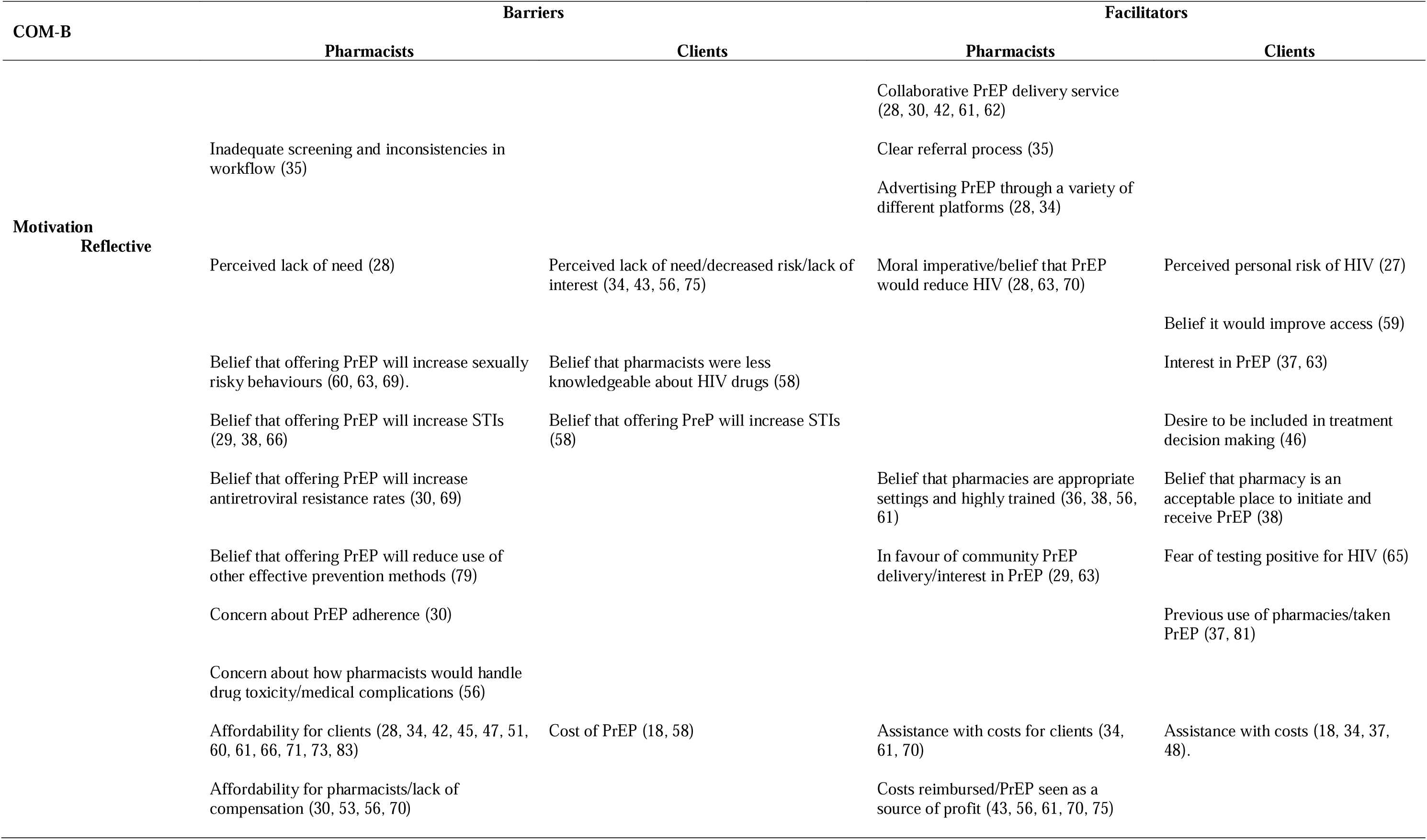
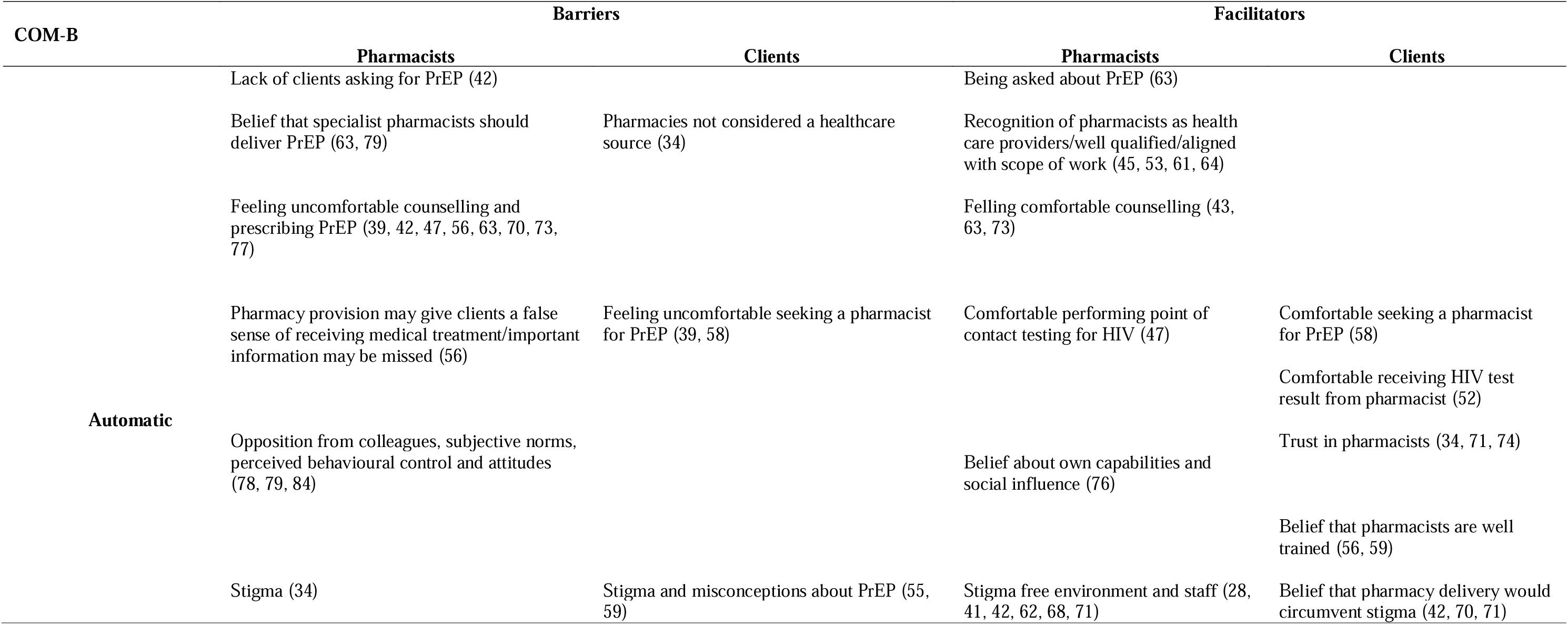
Summary of the barriers and facilitators of community pharmacy PrEP delivery for pharmacists and clients.

## Synthesis of results

### Pharmacist level barriers and facilitators

#### Capability barriers

Barriers to PrEP delivery for pharmacists included a lack of awareness among pharmacists and clients of the legislation authorising pharmacies in some states of America to provide PrEP (30, 36, 59). Other barriers included a lack of knowledge, skills and training among pharmacists about the medication, behaviour modification to reduce risks of acquiring HIV, prescribing PrEP and managing discussions about drug toxicity and/or medical complications (30, 38, 53, 56, 66, 67, 73). Additionally, a lack of guidelines on the circumstances around PrEP initiation and continuation (e.g., whether clients who self-test for HIV can be supplied PrEP) (67), lack of physician oversight when initiating PrEP from community pharmacies (18, 61), and a lack of clarity or knowledge about which clients would benefit from PrEP were reported to be barriers to identifying and counselling those at risk of HIV (42, 66, 71, 78, 79).

#### Opportunity barriers

Economic and social opportunity barriers for pharmacists included the country’s economic instability (60) and the inability to advertise branded medications in pharmacies (66). For the more specific aspects of PrEP delivery in community pharmacies, such as PrEP counselling and prescribing, pharmacists in the USA and Kenya reported the high turnover of staff (65) and the lack of skilled staff (e.g., only having one pharmacist on duty per shift) as barriers (28, 67, 70). Similarly, lack of staff time (28, 30, 47, 49, 53, 56, 70) was also found to be a barrier, particularly when needing longer consultation times (28) to initiate clients on PrEP (65) and carry out the required monitoring (e.g., kidney function) (33). There was also concern that an increased workload would negatively impact time allocated to delivery of other services (31). Pharmacists also identified physical barriers, such as the absence of private spaces or consultation rooms (53, 56, 61, 67, 70, 79), and laboratory facilities to carry out, process and record STI, HIV and kidney screening and monitoring (28, 56, 66, 67, 69–71).

#### Motivation barriers

Improving PrEP accessibility via community pharmacies was not judged to be a moral imperative by some pharmacists because pharmacy clients were not perceived to be at risk of acquiring HIV/HIV rates were assumed to be low (28). Other motivational barriers included experiencing opposition to PrEP delivery from colleagues (78, 79, 84), and concerns that increasing accessibility to PrEP would increase risky sexual behaviours (60, 63, 69), STIs (29, 38, 66), antiretroviral resistance (30, 69) and negatively impact the use of other available cost-effective STI preventative methods (e.g., condoms, abstinence) (79).

Pharmacists in the USA and Zimbabwe reported the affordability of PrEP, and whether medical insurance would cover the costs of counselling and prescribing (28, 34, 42, 45, 47, 51, 60, 61, 66, 71, 73, 83) to be significant barriers to PrEP delivery. Pharmacists were concerned that they (or the pharmacy business they worked for) would not profit or be adequately compensated for these services (30, 53, 56, 70). Pharmacists also reported concern about the stigma people could experience from community members when accessing PrEP (34), in addition to reporting a sense of discomfort discussing sexual health/histories, positive HIV test results, and counselling clients on behaviour modification and clinical trial data on PrEP effectiveness (39, 40, 47, 56, 63, 70, 73, 77). Some pharmacists also reported that they were reluctant to deliver PrEP without clients having had a prior medical consultation (61), and that PrEP delivery should be done by HIV speciality pharmacists only (63).

#### Capability facilitators

American pharmacists reported that having a good level of knowledge, and being able to identify who would benefit from PrEP (34, 71) would facilitate community pharmacy PrEP delivery. Other facilitators included pharmacists having received continuing education (CE) or specific HIV/PrEP education and/or training (29, 42, 64, 67, 79, 63, 66, 67, 77). Additional facilitators were identified to be pharmacists having the authority to be able to make recommendations about PrEP safety, efficacy and acquisition (36), having obtained a professional doctorate degree of pharmacy (PharmD) (63, 69, 76, 77), pharmacists having fewer years of pharmacy practice experience and having potentially been exposed to public health content during their education (42, 76, 77). Prior experience and familiarity of PrEP guidelines and subsequent confidence in prescribing PrEP (39, 69, 77) were also found to be facilitators.

#### Opportunity facilitators

Community pharmacies were seen as an ideal location for PrEP delivery. They were reported to be convenient in terms of location, opening hours and accessibility (39, 42, 47, 53, 56, 62, 63, 67). To overcome the barriers of staff capacity and time, pharmacists suggested using appointment systems to schedule PrEP screening activities (28, 49). Other suggestions included hiring more staff or having more staff on duty per shift to carry out PrEP counselling and dispensing (70). Pharmacists working in pharmacies with a larger number of full time staff were found to be more comfortable and able to spend the necessary amount of time counselling clients about PrEP (63). Pharmacists also suggested PrEP delivery could be facilitated by incorporating it into pre-existing services or pathways (e.g., STI testing) (56, 79).

Having a consultation room, private spaces or a separate room (28, 61, 66, 70) to have sensitive conversations in, in addition to facilities to enable STI and HIV testing (28), were reported to be facilitative of PrEP delivery. If there was a lack of facilities within the pharmacy, pharmacists reported that allowing clients to test themselves for HIV (65) or having a collaborative service with other health care providers could help facilitate PrEP delivery, adherence and continuation (28, 30, 42, 61, 62). Having a clear referral process for pharmacists to provide clients with the necessary treatment options for conditions identified during PrEP delivery was also found to facilitate PrEP delivery (35).

#### Motivation facilitators

Believing that pharmacists were health care providers (45, 53, 64), well qualified/trained, and pharmacies were appropriate settings to deliver PrEP (36, 38, 45, 56, 61) was found to facilitate PrEP delivery. Other motivational facilitators included pharmacists being in favour of community pharmacy PrEP delivery (29, 63), having an interest in HIV prevention (72), believing that PrEP could reduce the acquisition of HIV (28, 63, 70), feeling comfortable counselling about PrEP and HIV risk reduction behaviours (43, 63, 73) and performing point of contact testing (47).

To improve PrEP initiation, continuation and adherence, pharmacists reported that facilitating free access to PrEP or ensuring assistance with the cost (34, 61, 70) would facilitate delivery, particularly in the USA and for those most in need. Pharmacists also reported that the financial costs of PrEP to pharmacists would need to be reimbursed or covered by insurance companies (43, 56, 61, 70, 75) so that PrEP delivery was a source of income (70).

An additional facilitator of PrEP delivery for pharmacists was the belief that pharmacists should be able to provide a stigma free environment (28, 42, 62, 67, 71), that was culturally appropriate and convenient in which to have sensitive conversations (41).

### Client level barriers and facilitators

#### Capability barriers

For pharmacy clients, barriers to PrEP delivery via community pharmacies were reported to be a lack of awareness of PrEP, including its effectiveness (34, 49, 55, 57, 82). Clients also reported hesitancy toward pharmacy PrEP delivery because they lacked knowledge of pharmacists’ HIV and/or PrEP specialised knowledge/training (58).

#### Opportunity barriers

Actual or perceived environmental barriers to PrEP delivery in community pharmacy for clients included the lack of private consultation rooms (70, 81) or space to ensure privacy and confidentiality (34, 40). The location of the pharmacy (75) was also reported to be a barrier, particularly for clients relocating (43). Clients also recognised the lack of pharmacy facilities for laboratory testing (59) and pharmacist workloads as barriers to PrEP delivery (58).

#### Motivation barriers

Clients in the US were concerned about the cost of PrEP and pharmacists being unable to process their insurance (18, 58). Clients also reported concern that increasing access to PrEP could result in an increase in STIs (58). Additional barriers for clients were believing that there was a general and/or personal lack of need for PrEP (34, 43, 56, 75), believing pharmacist were not healthcare providers (34) and were not knowledgeable about HIV drugs (58) and that pharmacy PrEP delivery could result in medical errors (81). Clients also reported feeling less comfortable seeking a pharmacist for PrEP information and delivery (39, 58), preferring to speak to their physician (58), organisations that cater to the lesbian, gay, bisexual, transgender, queer/questioning and other (LGBTQ+) community or organisations that help cover the costs of PrEP (39). Some clients were also hesitant to be screened in pharmacies for fear of testing HIV-positive (65). Clients were also concerned about the misconceptions of PrEP being a medication for people living with HIV and that they could experience stigma from pharmacists (55, 59).

#### Capability Facilitators

Client awareness of PrEP was reported to facilitate PrEP delivery (31, 58, 59). Clients reported that the acceptability of community pharmacy PrEP delivery would be further facilitated if they were made aware of the regulatory policies (e.g., quality standards), and oversight (audits) that pharmacists had to adhere to (70), and any additional training or education for PrEP delivery that pharmacists had obtained (58, 70).

#### Opportunity Facilitators

Clients perceived the accessibility of community pharmacies in terms of location, speed of service and extended opening hours (42, 48, 56, 58, 62) to facilitate PrEP delivery, particularly if living in an urban area (48). Other important facilitators for clients were the presence of a private space to discuss and deliver PrEP to ensure confidentiality and respect (34, 40, 70), offering PrEP alongside other existing care pathways and services (e.g., opiate substitution therapy) and the presence of a pharmacist specialising in HIV service provision who could be easily identified (e.g., by their clothing) as the person to speak to about PrEP (40). Additional facilitators for clients included the pharmacies being able to provide HIV test results on the same day, offering ongoing monitoring for adherence support and risk reduction and being able to complete screening questionnaires prior to the consultation (46).

#### Motivation facilitators

Clients’ motivation to seek PrEP from community pharmacists was facilitated by interest in PrEP (37, 63), a desire to be involved in treatment decisions (46), a belief that pharmacy PrEP delivery would improve access to PrEP (34, 71, 74), feeling comfortable seeking PrEP information and prescriptions from pharmacists (58), receiving HIV test results from pharmacists (38, 56) and having trust in pharmacists (42, 70, 71). Clients who perceived themselves to be at a greater risk of HIV due to engagement in condomless sex or sex with multiple partners, were also found to be more willing to be screened in community pharmacies (27).

Believing that pharmacists were well trained to deliver PrEP (56) and that pharmacies were acceptable places to initiate and receive PrEP was also found to facilitate PrEP delivery (38, 56, 59). Other facilitators included PrEP being available free or at a low cost to clients (18, 34, 37, 48, 65), having insurance to cover the costs of PrEP (27), and believing that PrEP delivery in community pharmacies could circumvent the stigma associated with PrEP and HIV, in part by offering clients the opportunity to obtain PrEP discretely (42, 70, 71).

## Discussion

In this scoping review we mapped the barriers and facilitators of community pharmacy PrEP delivery to the COM-B model. The current review highlights the increase in research in this area, particularly in the USA, shortly after pharmacists in the USA were authorised to deliver PrEP. Whilst there have been systematic and scoping reviews on pharmacy PrEP delivery (53, 80) none of the reviews have aimed to identify and map the potential barriers and facilitators of PrEP delivery, according to a behavioural theory or model. Subsequently, the findings from the current review, provide important insight for the development of targeted interventions and strategies to optimise potential community pharmacy PrEP delivery. In this review, barriers identified included lack of PrEP awareness, familiarity knowledge, skills and training (capability), lack of staff capacity, time and pharmacy facilities (opportunity), the financial cost of PrEP to pharmacists and clients and the belief that PrEP delivery could lead to risky behaviours and higher rates of STIs (motivation). Facilitators identified included improving client and pharmacist awareness of PrEP, and the provision of PrEP training and education (capability), having an appointment system, using pre-existing pathways or services the accessibility of community pharmacies (opportunity) and the belief that PrEP delivery could be a source of income or profit and contribute to reductions in local and population level HIV rates (motivation).

In translating the findings to community pharmacy PrEP delivery in the UK, some of the opportunity barriers presented have already been addressed. The UK vision for community pharmacy is for more integration into the NHS, to provide more clinical services, to support and manage demands on the NHS (85). Consequently, most pharmacies in the UK now have private consultation rooms and in some regions, there are already well-established links with other health care providers (e.g., sexual health clinics) for the provision of services. For example, the emergency contraception referral pathway is supported by regular training and collaboration with Local Pharmaceutical Committees (LPCs). Some of these services could offer potential opportunities for expansion to include the provision of PrEP, particularly the reproductive and sexual health services that are requested most frequently (e.g., condoms, STI self-testing kits, emergency contraception) (86). Patient Group Directions (PGDs) which authorise pharmacy professionals to supply particular medications to clients presenting in person, also offer potential opportunities for PrEP delivery in the UK.

Other barriers identified, such as the lack of skilled staff and staff capacity highlight the increasing workload of pharmacists (13, 14, 87) and the potential lack of feasibility of introducing PrEP delivery to community pharmacies (88). Although hiring more staff was identified to help overcome staff capacity barriers, results from the Pressures Survey conducted by the Pharmaceutical Services Negotiating Committee (PSNC) found 71% of community pharmacies were experiencing a shortage of pharmacists (89). Consequently, there appears to be a lack of pharmacists to employ even though there is demand. A better understanding of current issues facing pharmacists in the UK could improve awareness and inform future investment in education and training in addition to effective workplace planning for pharmacy service provision, including PrEP delivery.

The accessible location, and convenient walk-in nature of pharmacies, as highlighted in this review, could offer an easy, less intimidating environment from which to access PrEP. This may be particularly important for individuals with lower levels of literacy who find the increasing use of digital technology required to access health services challenging (90).

Community pharmacy PrEP delivery could therefore help to reduce inequalities amongst those who are disproportionately affected by HIV, in part because ease of access has been shown to be key to the uptake and maintenance of PrEP (80, 91). Similarly, addressing the motivational barriers identified by pharmacists and clients could help circumvent any stigma, facilitate awareness of LGBTQ+ issues, and improve client confidence in pharmacists’ medical expertise and capability to provide services. Future research needs to examine the perceived accessibility and feasibility of UK pharmacies for PrEP delivery, particularly among pharmacists and individuals at elevated risk of acquiring HIV who are not currently accessing PrEP from sexual health clinics. This research should also explore whether the stigmatising attitudes of some pharmacists could act as barriers to service provision (13).

Overall, the results suggest that in order to change behaviour and facilitate community pharmacy PrEP delivery in the UK, pharmacists and clients would need to be provided with a training intervention that fosters beliefs about the positive impact of PrEP delivery, stimulates and harnesses client’s interest and provides financial support for pharmacists and clients to enhance motivation for PrEP delivery, initiation, continuation and adherence.

There are limitations to this scoping review. The majority of the research reviewed was from the USA so the barriers and facilitators presented may be specific to this population (e.g. payment for healthcare and differing contractual frameworks). More work is needed to understand UK-specific barriers and facilitators. Further, a number of studies included pharmacists working in settings other than community pharmacies (e.g., GP practices and hospitals). This made the findings specific to community pharmacists hard to distinguish.

Notwithstanding this, the current scoping review has methodological strengths (i.e., searching of multiple databases, handsearching of citations) and employs the COM-B model as a framework to synthesize the evidence.

## Conclusions

This scoping review provides valuable insights into the barriers and facilitators of PrEP delivery in community pharmacies. It highlights the multifaceted nature of PrEP delivery and underscores the need to consider individual external and reflective factors influencing the behaviour of pharmacists and clients in providing and accessing PrEP services. This review is the first step toward developing theory and an evidence-based intervention that is acceptable and effective for UK community pharmacies. By taking a comprehensive approach that considers all aspects of the COM-B, community pharmacies in the UK could become crucial players in expanding PrEP accessibility and uptake, contributing significantly to HIV prevention efforts.

## Competing interests

None to declare

## Authors contributions

CH, JK, SD, HF, JS, CB, JC, LH, SC and JH contributed to the research planning. AS & SD contributing to the literature search, AS, JH, & CH to study selection and CH, JH, JK, HF, SD to data extraction. CH and JH drafted all versions of the manuscript, and all authors contributed to and approved the final version for publication.

## Funding

This research was funded by Gilead Sciences, Inc, and supported by the National Institute for Health and Care Research Applied Research Collaboration West (NIHR ARC West) and the National Institute for Health and Care Research Health Protection Research Unit (HPRU) in Behavioural Science and Evaluation at the University of Bristol, and in Sexually Transmitted and Blood Borne Infections at UCL, both in partnership with UKHSA.. The views expressed in this article are those of the authors and not necessarily those of the NIHR, UKHSA or the Department of Health and Social Care.

## Data availability

Data is available upon reasonable request from the corresponding author.

## Supporting information

Supplementary Table 1. Search terms and strategies used for the five main bibliographic databases and give review databases

Supplementary Table 2. Summary of the total (N=56) included literature, methodological characteristics, study objectives and population

## Notes

### Competing Interest Statement

The authors have declared no competing interest.

