## Supplementary Table 1 for "Facilitators and barriers to community pharmacy PrEP delivery: A scoping review"

Search terms and strategies used for the five main bibliographic databases and five review databases.

| | |
|---|---|
| | **Ovid MEDLINE(R) ALL** |
| 1 | exp HIV/ |
| 2 | exp HIV Infections/ |
| 3 | (HIV or HIV-1* or HIV1* or HIV-2* or HIV2* or human immunodeficien* vir* or human immunedeficien* vir* or human immun* deficien* vir* or acquired immunodeficien* syndrom* or acquired immunedeficien* syndrome* or immun* deficien* syndrom*).mp. |
| 4 | AIDS related.mp. |
| 5 | or/1-4 |
| 6 | Pre-Exposure Prophylaxis/ |
| 7 | (PrEP or pre expos* prophyla* or preexpos* prophyla*).mp. |
| 8 | *Anti-Retroviral Agents/tu and (prevent* or prophyla*).af. |
| 9 | ((antiretrovir* or anti-retrovir* or HIV) adj prophyla*).mp. |
| 10 | (tenofovir or TNF or TDF or PMPA or viread or emtricitabine or EMC or truvada or emtriva or coviracil).mp. |
| 11 | impact trial.mp. |
| 12 | or/6-11 |
| 13 | 5 and 12 |
| 14 | Community Pharmacy Services/ or *Pharmacy/ |
| 15 | (pharmacy or pharmacies or pharmacist?).ti,kf |
| 16 | ((communit* or local* or retail or "in store" or supermarket* or walk-in or drop-in or pop-in or pop-up) adj4 (pharmacy or pharmacies or pharmacist?)).tw. |
| 17 | ((pharmacy or pharmacies or pharmacist?) adj4 (based or deliver* or intervention* or located or location? or led or program* or run or set or setting?)).tw. |
| 18 | or/14-17 |
| 19 | 13 and 18 |
| 20 | (PrEP or pre expos* prophyla* or preexpos* prophyla*).ti. |
| 21 | 18 and 20 |
| 22 | 19 or 21 |
| | **Ovid Embase** |
| 1 | exp Human immunodeficiency virus/ |
| 2 | exp Human immunodeficiency virus infection/ |
| 3 | (HIV or HIV-1* or HIV1* or HIV-2* or HIV2* or human immunodeficien* vir* or human immunedeficien* vir* or human immun* deficien* vir* or acquired immunodeficien* syndrom* or acquired immunedeficien* syndrome* or immun* deficien* syndrom*).tw,kf. |
| 4 | AIDS related.tw,kf. |
| 5 | or/1-4 |
| 6 | Pre-exposure prophylaxis/ |
| 7 | (PrEP or pre expos* prophyla* or preexpos* prophyla*).tw,kf. |
| 8 | ((antiretrovir* or anti-retrovir* or HIV) adj prophyla*).tw,kf. |
| 9 | tenofovir.hw. or (tenofovir or TNF or TDF or PMPA or viread or emtricitabine or EMC or truvada or emtriva or coviracil).tw,kf. |
| 10 | impact trial.mp. |
| 11 | or/6-10 |
| 12 | 5 and 11 |
| 13 | "pharmacy (shop)"/ |
| 14 | (pharmacy or pharmacies or pharmacist?).ti,kf. |

| | |
|---|---|
| 15 | ((communit* or local* or retail or "in store" or supermarket* or walk-in or drop-in or pop-in or pop-up) adj4 (pharmacy or pharmacies or pharmacist?)).tw. |
| 16 | ((pharmacy or pharmacies or pharmacist?) adj4 (based or deliver* or intervention* or located or location? or led or program* or run or set or setting?)).tw. |
| 17 | or/13-16 |
| 18 | 12 and 17 |
| 19 | (PrEP or pre expos* prophyla* or preexpos* prophyla*).ti. |
| 20 | 17 and 19 |
| 21 | 18 or 20 |
| 22 | |
| | **Ovid APA PsycInfo** |
| 1 | exp Pre-Exposure Prophylaxis/ |
| 2 | (PrEP or pre expos* prophyla* or preexpos* prophyla*).tw,id. |
| 3 | ((antiretrovir* or anti-retrovir* or HIV) adj prophyla*).tw,id. |
| 4 | (tenofovir or TNF or TDF or PMPA or viread or emtricitabine or EMC or truvada or emtriva or coviracil).tw,id. |
| 5 | or/1-4 |
| 6 | pharmacists/ or pharmacy/ |
| 7 | (pharmacy or pharmacies or pharmacist?).tw,id. |
| 8 | 6 or 7 |
| 9 | 5 and 8 |
| | **Cochrane Central Register of Controlled Trials** (CENTRAL) |
| 1 | PrEP OR ((pre expos* or preexpos*) NEAR/2 prophyla*) OR ((antiretrovir* or anti-retrovir* or HIV) NEAR/2 prophyla*) OR tenofovir or TNF or TDF or PMPA or viread or emtricitabine or EMC or truvada or emtriva or coviracil):ti,ab,kw |
| 2 | ((communit* or local* or retail or "in store" or supermarket* or shop or walk-in or drop-in or pop-in or pop-up) NEAR (pharmacy or pharmacies or pharmacist*)):ti,ab,kw |
| 3 | ((pharmacy or pharmacies or pharmacist*) NEAR (based or deliver* or intervention* or located or location* or led or program* or run or set or setting*)):ti,ab,kw |
| 4 | (#1 AND (#2 OR #3)) |
| 5 | ((PrEP OR ((pre expos* or preexpos*) NEAR/2 prophyla*) OR ((antiretrovir* or anti-retrovir* or HIV) NEAR/2 prophyla*) OR tenofovir or TNF or TDF or PMPA or viread or emtricitabine or EMC or truvada or emtriva or coviracil) AND (pharmacy or pharmacies or pharmacist*)):ti |
| 6 | ((PrEP OR ((pre expos* or preexpos*) NEAR/2 prophyla*) OR ((antiretrovir* or anti-retrovir* or HIV) NEAR/2 prophyla*) OR tenofovir or TNF or TDF or PMPA or viread or emtricitabine or EMC or truvada or emtriva or coviracil) NEAR/10 (pharmacy or pharmacies or pharmacist*)):ab |
| 7 | (#4 OR #5 OR #6) |
| | **Cumulative Index to Nursing and Allied Health Literature** (CINAHL) (EBSCOHost) |
| 1 | TI ( PrEP or ((pre-expos* or preexpos*) AND prophyla*) ) OR AB ( PrEP or ((pre-expos* or preexpos*) N2 prophyla*) ) |
| 2 | TI ( ((antiretrovir* or anti-retrovir* or HIV) AND prophyla*) ) OR AB ( ((antiretrovir* or anti-retrovir* or HIV) W1 prophyla*) ) |
| 3 | TI ( (tenofovir or TNF or TDF or PMPA or viread or emtricitabine or EMC or truvada or emtriva or coviracil) ) OR AB ( (tenofovir or TNF or TDF or PMPA or viread or emtricitabine or EMC or truvada or emtriva or coviracil) ) |
| 4 | (S1 OR S2 OR S3) |
| 5 | (MH "Pharmacists") |
| 6 | TI ( (pharmacy or pharmacies or pharmacist*) ) OR AB ( (pharmacy or pharmacies or pharmacist*) ) |

| | |
|---|---|
| 7 | (S5 OR S6) |
| 8 | (S4 AND S7) |
| 9 | S4 AND S7) Limiters - Exclude MEDLINE records |
| | The following review databases were also searched, using a pragmatic search for PrEP: (*HIV* AND (*PrEP* or "*pre-exposure prophylaxis*") AND (*pharmacy* OR *pharmacies* OR *pharmacists*)) Cochrane Database of Systematic Reviews (CDSR) (0); the Database of Promoting Health Effectiveness Reviews (DoPHER) (0); Epistemonikos (55) (10 selected: 5 duplicates, 5 new); Health Evidence (1 (irrelevant)); NIHR Health Technology Assessments (0). |
