## Supplementary Table 2 for "Facilitators and barriers to community pharmacy PrEP delivery: A scoping review"

*Summary of the total (N=56) included literature, the methodological characteristics, study objectives and population*

| Author | Year | Location | Publication type | Study design | Study objective | Study population |
|---|---|---|---|---|---|---|
| Alohan et al (27) | 2023 | USA | Original research | Cross sectional survey | To examine the correlates of patient (men-who-have-sex-with-men, MSM) willingness to be screened for PrEP in pharmacies | Customers (MSM) |
| Bellman et al.(28) | 2022 | USA | Original research | Cross sectional survey and interviews | To access implementation of PrEP in San Francisco pharmacies and barriers and successes of implementation | Pharmacists |
| Bouetard & Cordel (29) | 2021 | France | Conference abstract | Cross sectional survey | To measure community pharmacists' knowledge, experience and perception regarding PrEP | Pharmacists |
| Broekhuis et al. (30) | 2018 | USA | Original research | Cross sectional survey | To characterise pharmacists' knowledge about and willingness to provide PrEP | Pharmacists |
| Booker et al. (31) | 2023 | USA | Original research | Cross sectional survey and interviews | To determine pharmacists' acceptance of a pharmacist PrEP prescribing service | Pharmacists |
| Bruno & Saberi (32) | 2012 | USA | Commentary | | To discuss reasons for a protocol-based, clinical pharmacist-run PrEP clinic. | N/A* |
| Burns et al. (33) | 2023 | USA | Original research | Cross sectional survey | To determine the perceived feasibility and acceptability of prescribing PrEP by pharmacists | Pharmacists* |
| Cernasev et al. (34) | 2022 | USA | Original research | Focus groups | To characterise Tennessee pharmacists' perceptions about access to PrEP | Pharmacists |
| Cernasev et al. (35) | 2023 | Only studies conducted in the USA | Review | Systematic review | To explore the tole of pharmacists, pharmacy services and interpersonal collaborations for persons seeking PrEP | |
| Clauson et al. (36) | 2009 | USA | Commentary | N/A | To discuss reasons for a protocol-based, clinical pharmacist-run PrEP clinic. | N/A |

| Author | Year | Location | Publication type | Study design | Study objective | Study population |
|---|---|---|---|---|---|---|
| Crawford et al. (37) | 2021 | USA | Original research | Cross sectional survey | To examine the correlates of willingness to discuss PrEP with pharmacy staff and screen for PrEP in a pharmacy setting. MSM. | Customers (MSM) |
| Crawford et al. (38) | 2021 | Only studies conducted in USA | Review | Systematic literature review | To synthesise the recent literature on the role of pharmacists across the HIV prevention and care continuums | |
| Crawford et al. (39) | 2020 | USA | Original research | Qualitative interviews | To understand the perceptions and support for pharmacy-based pre-exposure prophylaxis (PrEP) delivery among pharmacists and MSM | Pharmacists and customers (MSM) |
| Crawford et al. (40) | 2022 | USA | Original research | Pilot of protocol | To measure feasibility, acceptance, safety, and PrEP use at baseline and at 3-months | Customers (MSM) |
| Dong et al. (41) | 2019 | USA | Opinion | N/A | To respond to Havens et al 2019 study (below) and provide an alternative perspective, with examples of how PrEP has been successfully implemented in pharmacy settings. | N/A |
| Farmer et al. (42) | 2019 | USA | Review | Literature review | To summarise evidence on pharmacist involvement in different models of care providing PrEP services and to identify opportunities to maximise and expand the role of the pharmacist to improve access to PrEP. | |
| Garrison et al. (43) | 2021 | USA | Review | Narrative review | To summarise interventions (2017-2020) aimed at improving PrEP uptake, adherence and persistence of PrEP. | |
| Gregory (44) | 2020 | USA | Commentary | N/A | To report on the new law in California that allows pharmacists to provide PrEP | N/A |
| Griffith et al. (45) | 2022 | USA | Commentary | USA | To discuss access and adherence to PrEP in community pharmacy? | N/A |

| Author | Year | Location | Publication type | Study design | Study objective | Study population |
|---|---|---|---|---|---|---|
| Goswami (46) | 2022 | USA | Original research (PhD dissertation) | Survey and discreet choice experiment | To elicit preferences for attributes of a community pharmacy-based PrEP delivery program | Customers (MSM) |
| Havens et al.(47) | 2019 | USA | Original research | Quantitative analysis of use of service/feasibility trial | To assess the acceptability and feasibility of a pharmacist-led HIV screening and PrEP program (P-PrEP) for individuals at risk for HIV acquisition in Omaha, Nebraska | Customers |
| Hazen et al. (48) | 2017 | USA | Conference abstract | Retrospective cohort | To characterise PrEP medication utilisation patterns and factors associated with medication adherence | Customers |
| Hopkins et al.(49) | 2021 | USA | Original research | Qualitative Interviews | To assess pharmacists' and pharmacy technicians' perspectives regarding the implementation of PrEP screening and dispensing | Pharmacy technicians and pharmacists |
| Huang et al (50) | 2022 | USA | Original research | Cross sectional analysis of national pharmacy database | To better understand PrEP cessation | Pharmacy database |
| Hughes et al. (51) | 2019 | Canada | Guidelines | N/A | To provide an overview of guidelines and highlight the role pharmacists can have in HIV prevention | N/A |
| Josma et al. (52) | 2023 | USA | Original research | Qualitative Interviews | To examine black men-who-have-sex-with-men (BMSM) beliefs about accessing PrEP in pharmacies | Customers (BMSM) |
| Kazi et al (18) | 2019 | USA | Commentary | | To discuss pharmacy led PrEP delivery | |
| Kennedy et al. (53) | 2022 | Mixed | Review | Systematic review | To evaluate the evidence for distributing PrEP through pharmacies. | Pharmacists and people interested in PrEP |
| Khosropour et al. (54) | 2020 | USA | Original research | Pilot study | Pilot to facilitate PrEP uptake and decrease time to PrEP initiation | Customers |
| Khosropour et al. (55) | 2023 | USA | Original research | Mixed methods evaluation | To describe PrEP initiation and persistence of individuals who participated in a community | Customers |

| Author | Year | Location | Publication type | Study design | Study objective | Study population |
|---|---|---|---|---|---|---|
| | | | | | pharmacy PrEP delivery program and to examine the barriers motivations and facilitators to PrEP initiation and persistence | |
| Koester et al. (56) | 2020 | USA | Original research | Qualitative case study | To assess attitudes among key stakeholders about a California policy to allow community pharmacists to deliver HIV PrEP. | Pharmacists and stakeholders |
| Lopez et al. (57) | 2020 | USA | Commentary | N/A | To encourage key stakeholders to advocate for implementing community pharmacy–initiated PrEP and provide recommendations for implementing PrEP in a community pharmacy. | N/A |
| Lutz et al. (58) | 2021 | USA | Original research | Cross sectional survey | To assess patient perspectives of pharmacist PrEP prescribing and identify potential barriers to acceptance of pharmacy prescribed PrEP | Patients receiving antiretroviral medications |
| MacDonald et al. (59) | 2023 | USA | Original research | Cross sectional survey and interviews | To determine target users acceptance of a PrEP prescribing service by pharmacists | Customers |
| Matyanga et al. (60) | 2014 | Zimbabwe | Original research | Cross sectional survey | To assess pharmacists' knowledge, perception and willingness to provide PrEP. | Pharmacists |
| Mayers et al. (61) | 2018 | USA | Review | Critical review | To analyse the current state of PrEP implementation in the US by reviewing barriers and innovative solutions to enhance PrEP access and uptake | |
| McCree et al. (62) | 2020 | USA | Report | | To discuss potential roles that pharmacists and pharmacies can play in delivering PrEP | N/A |
| Meyerson et al.(63) | 2019 | USA | Original research | Cross sectional survey | To identify factors associated with PrEP dispensing and comfort with PrEP counselling among community pharmacists to inform the design of evidence based pharmacy-practice interventions | Pharmacists (managing pharmacists) |

| Author | Year | Location | Publication type | Study design | Study objective | Study population |
|---|---|---|---|---|---|---|
| Myers et al. (64) | 2019 | USA | Commentary | | To share recommendations on community pharmacy PrEP delivery | N/A |
| Nakambale et al. (65) | 2023 | Kenya | Original research | Observations | To identify early implementation barriers of pharmacy based PrEP delivery and actions that pharmacy providers took to help address these | Pharmacists and customers |
| Okoro & Hillman (66) | 2018 | USA | Original research | Cross sectional survey | To assess knowledge and experience, describe perceptions and attitudes and identify training needs of community based pharmacists | Pharmacists |
| Ortblad et al. (67) | 2020 | Kenya | Original research | Stakeholder consultation* | To develop a pathway for pharmacy-based PrEP delivery in Kenya | PrEP stakeholders |
| Ortblad et al.(68) | 2020 | Kenya | Commentary | | To discuss the adoption of pharmacy- based PrEP delivery in Africa in relation to the USA | N/A |
| Przybyla et al.(69) | 2019 | USA | Original research | Cross sectional survey | To assess pharmacy students awareness, knowledge and perceptions towards HIV, PrEP, confidence and intentions to counsel customers on PrEP and preferred PrEP training | Pharmacy students enrolled in a doctor of pharmacy (PharmD) |
| Roche et al.(70) | 2021 | Kenya | Original research | Qualitative Interviews | To identify factors that may influence the implementation of pharmacy-based PrEP delivery in Kenya, to inform the design and implementation of a pharmacy PrEP care pathway. | pharmacy clients. pharmacy providers, PrEP clients, PrEP providers |
| Rousseau et al. (71) | 2021 | Mixed | Review | | To summarise novel platforms for HIV prevention outside of the traditional health facilities environment | |

| Author | Year | Location | Publication type | Study design | Study objective | Study population |
|---|---|---|---|---|---|---|
| Sawkin et al. (72) | 2016 | USA | Conference abstract | Unclear | To develop and test a pharmacy led clnic to expand access to PrEP | Unclear |
| Shaeer et al. (73) | 2013 | USA | Conference abstract | Cross sectional survey | To assess pharmacists experience with, knowledge of, and perceptions about HIV PrEP and identify areas for pharmacist training. | Pharmacists |
| Smith et al. (74) | 2021 | UK | Original research | Qualitative Interviews | To explore views of people who inject drugs (PWID) who might benefit from PrEP provision and service providers working with PWID to understand willingness to use PrEP and literacy of PrEP, contributing to the development of a PrEP service | Customers (PWID) |
| Tung et al. (75) | 2019 | USA | Original research | Service evaluation | To describe the service (pharmacist managed PrEP clinic in a community pharmacy setting) and report initial experiences | Pharmacists and customers |
| Unni et al. (76) | 2016 | USA | Original research | Cross sectional survey | To measure pharmacists knowledge about PrEP, perceptions about their PrEP knowledge and intentions to counsel customers about PrEP | Pharmacists |
| Wilby et al. (77) | 2020 | Mixed | Review | Narrative review | To identify priority areas and key gaps for continuing professional development (CPD) needs relating to PrEP for practicing pharmacists | |
| Yoong et al. (78) | 2016 | Canada | Original research | Cross sectional survey | Evaluate pharmacists attitudes and readiness for PrEP | Pharmacists |

| Author | Year | Location | Publication type | Study design | Study objective | Study population |
|---|---|---|---|---|---|---|
| Yoong et al. (79) | 2013 | Canada | Conference abstract | | To determine Canadian pharmacists enthusiasm for PrEP and their opinions on PrEP implementation. | Pharmacists |
| Zhao et al. (80) | 2022 | USA | Review | Scoping review | To explore pharmacy based initiatives to increase PrEP use | |
| Zhu et al. (81) | 2020 | USA | Original research | Cross sectional survey | To determine customers perceptions of pharmacist-prescribed PrEP, determining whether perceptions differed between customers who had previously been prescribed PrEP and those who had not and those who had at least one indication for PrEP verses those who did not. To identify what customer characteristics make customers more willing to seek PrEP from pharmacies. | Customers |

*Note*: *Record does not include or refer to community based pharmacists only, clinical pharmacists or other health care professionals are included or referred to.
